# IDENTIFICATION OF GENOTYPE III OF HEPATITIS DELTA VIRUS IN ANDEAN AND AMAZONIAN COMMUNITIES OF PERU

**DOI:** 10.1101/2022.05.03.22274000

**Authors:** Johanna N. Balbuena-Torres, Lorena Santos-Solis, Ronald D. Navarro-Oviedo, Cesar Cabezas

## Abstract

**Objectives:** to identify the genotypes of Hepatitis Delta Virus (HDV) circulating in populations of the inter-Andean valley of Huanta and three indigenous peoples of the Peruvian Amazon.

**Materials and Methods:** Observational and cross-sectional study, from 582 reactive samples for anti-HBc-HBV antibodies in inhabitants of the andean province of Huanta (Ayacucho) and the Amazonian towns of Matsés, Kandozi and Chapra (Loreto). Analysis was performed for HDV infection markers: anti-HDV IgM and anti-HDV IgG by ELISA using Wantai brand kits. Anti-HDV positive samples by ELISA were processed with the nRT-PCR method for the detection of HDV RNA. HDV genotype was determined by direct Sanger-type sequencing and phylogenetic analysis of the R0 fragment. 111 reference sequences from GenBank were used. The 42 sequences of the study were edited, assembled and cut with the programs Chromas 2.6.5, Bioedit v7.2, ClustalW v.1.6 of Mega v.7.0 and the Gblocks server. Phylogenetic and evolutionary analysis was performed with the following software: Beast V2.5.2, Jmodeltest v2.1.10, Tracer v1.7.1, Tree Annotator and Figtree v1.4.4. The Bayesian Yule and Birth Death skyline serial models were used, the MCMC at 30 and 80 million respectively, with the relaxed uncorrelated Exponential molecular clock. Summary and central tendency measures were calculated using the program in STATA 14.0.

**Results:** The mean age was 38 years (0 to 86 years), 52.75% (N=307) were women. 582 blood samples positive for anti-HBc were analyzed for anti-HVD antibodies using the ELISA method, with 101 positive samples being found. HDV RNA was detected in 49.50% of the anti-HDV ELISA reactive samples. Phylogenetic analysis determined the presence of genotype 3.

**Conclusions:** The presence of HDV genotype 3 in Andean and Amazonian communities of Peru is evidenced.

## INTRODUCTION

It is estimated that there are between 15 and 20 million people infected with the hepatitis delta virus (HDV) worldwide, which corresponds to 5% of those infected with HBV^1^. This infection can occur as an HBV/HDV superinfection or co-infection. Chronic HBV/HDV infection can cause more severe forms of the disease with further progression to liver fibrosis and hepatocellular carcinoma (HCC) ^2^.

Hepatitis delta virus (HDV) is a defective, covalently closed, circular single-stranded RNA virus that requires the hepatitis B virus (HBV) surface antigen (HBsAg) to infect and replicate in hepatocytes. 1679 to 1697 nucleotides, with a single ORF that encodes two isoforms of the Delta Antigen (HDAg) protein: HDAg-L and HDAg-S^3^ that differ in the additional 19 amino acids (aac.) at the carboxyl-terminal end of HDAg-L^4^. The HDAg-S isoform promotes viral replication and HDAg-L participates in the assembly of the virion^5^.

HDV is classified into 8 genotypes whose intergenotypic divergence is between 35% and 40% while intragenotypic heterogeneity is <20%. The geographical distribution of these genotypes is varied: VHD-1 has a worldwide distribution and the other genotypes are specific to a geographical region, VHD-2 and VHD-4 are prevalent in Japan (Asia), VHD-3 in Latin America mainly in the Amazon and VHD-5 to VHD-8 in Africa^6^

The prevalence of HDV infection varies according to geographical area, with Africa, the Mediterranean, Asia, Japan, Taiwan, Pakistan, Middle East and South America being considered as the areas with the highest prevalence. South America, in the last 3 decades, has reported a prevalence of 22.37%, however, it must be taken into account that most of the studies have been carried out in areas considered endemic and with geographic isolation^7^.

In Peru, there are some studies on the prevalence of HDV which indicate that the native communities of the Amazon and the Andean areas such as Abancay, Ayacucho and Andahuaylas are the areas with the highest prevalence. A study carried out in 8 localities of the Pampas River (Ayacucho-Andahuaylas), in HBsAg-positive schoolchildren, determined a prevalence of 16.7% for VHD^8^, while in the city of Huanta, 14.7% of anti-HBc-positive schoolchildren had HDV^9^. In HBsAg-positive Amazonian inhabitants, the prevalence was 39% ^10^ and in Abancay inhabitants with HBV infection it was 9%^11^. In 1992-1993, in an outbreak in soldiers from the Amazon jungle of Peru with a clinical diagnosis of hepatitis, the presence of HDV was determined in 64% of them, and it was also determined that they corresponded to genotype III^12^

Current studies indicate that the prevalence of HDV has decreased in recent years, going from an area of high endemicity to an area of low endemicity. The HBV vaccination campaigns carried out by MINSA and the screening for HBV have contributed to the decrease in the prevalence of HDV in the regions of greatest endemicity.

This study has been carried out to identify the HDV genotype that is circulating in Andean communities such as Huanta (Ayacucho) and in Amazonian indigenous peoples: Kandozi, Chapra and Matsés (Loreto). The range of nucleotide substitution, the population dynamics and the time of the most recent common ancestor (tMRCA) have been estimated.

## MATERIALS AND METHODS

Population and sample size an observational, analytical and descriptive cross-sectional cohort study was carried out, with 582 samples positive for HBV Anti-HBc. All were determined by the ELISA method. These samples came from three native Amazonian communities Matsés, Kandozi and Chapra of the Loreto Region; and the inter-Andean valley of Huanta located in the Ayacucho Region. The blood samples were collected during the years 2010 to 2012 and stored in refrigeration in the Laboratory of Hepatitis and Enterovirus-INS.

The three Amazonian native communities are located in the department of Loreto. The Kandozi and Chapra are located in the province of Datem del Marañón, in the Pastaza and Morona basin. The Matsés are located in the province of Requena, on the banks of the Gálvez and Yaquirana rivers and the Añushiyacu stream. The province of Huanta, in the Ayacucho region, is an inter-Andean valley located on the eastern slope of the Andes mountain range in Peru. (Figure No. 1)

**Figure N° 1.**
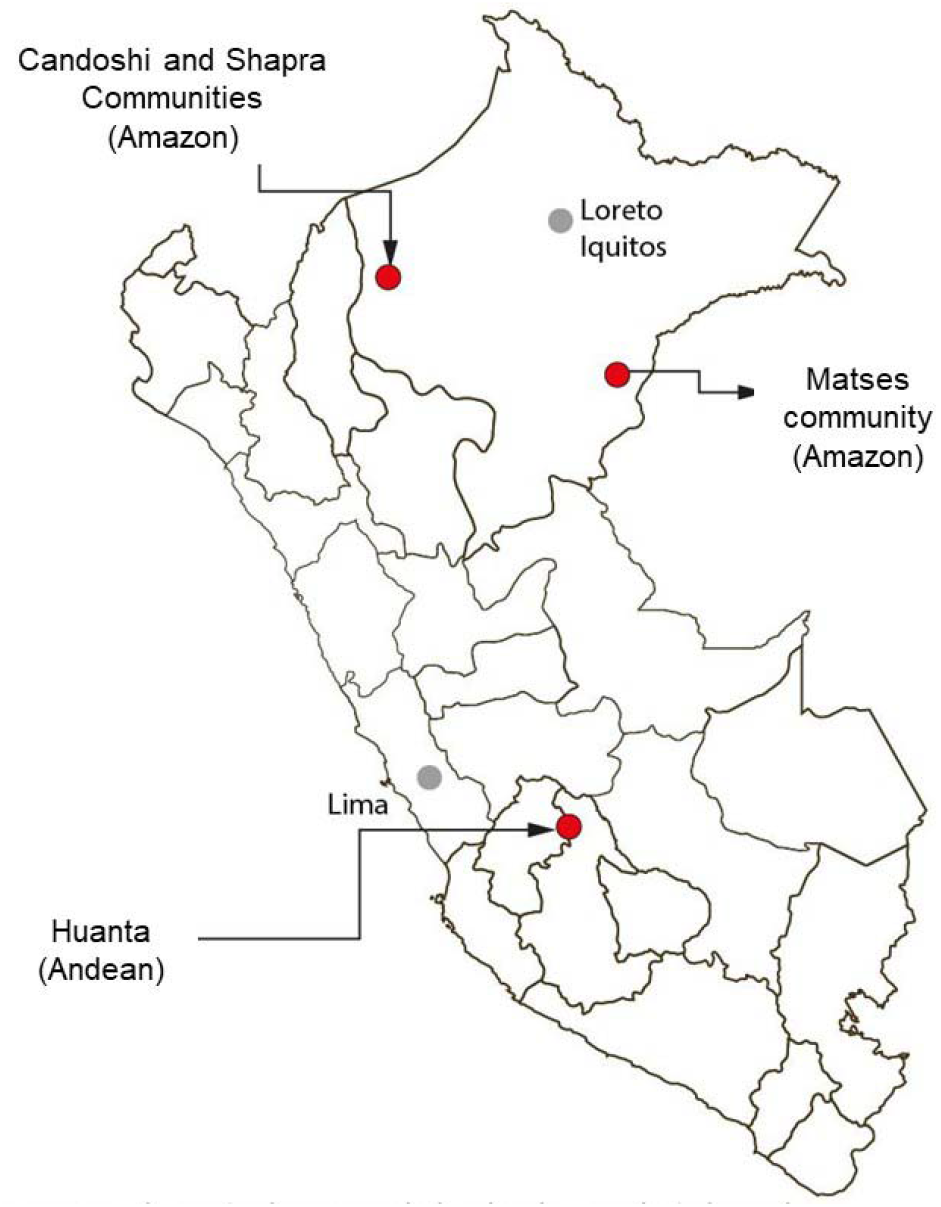
Location of communities where the study was conducted

### Laboratory analysis

#### Serology and viral load

In the facilities of the Hepatitis Laboratory, laboratory analyzes were carried out to determine the IgG or IgM serological markers of HDV using Wantai ELISA tests (Beijing Wantai Biological Pharmacy) and the Robonik ELISA washer and reader. All analyzes were performed following the manufacturer’s instructions.

For samples from chronic HBV carriers, HBV viral load was determined by quantitative real-time polymerase chain reaction (qPCR) using the COBAS AmpliPrep/COBAS TaqMan HBV Test Kits, version 2.0 (Roche Molecular Diagnostics, Branchburg, New Jersey), whose lower limit of detection is 20 IU/mL.

### HDV RNA extraction and nRT-PCR

RNA was extracted from HDV IgG or IgM positive samples using the commercial QIAamp viral RNA kit (Qiagen, Germany) and 140 µl of serum. The extracted RNA was denatured at 95°C for 5 minutes together with the random oligonucleotides, 12 µL of the denatured RNA was added to the reverse transcription mixture for cDNA synthesis (RT-PCR) using the High-Capacity cDNA Reverse Transcription kit (Applied Biosystems-Life Technologies, Carlsbad, CA). 10 µL cDNA, with 40 µL reaction mix [1x buffer, 1.5 mM MgCl2, 0.2 of each dNTPs, 0.5 pmol/µL of each 853IU-1302OD oligonucleotide, and 0.025 U/µL of Taq DNA polymerase (Invitrogen). For the nested PCR, 5 µl of the amplified primer and 45 µl of the PCR mixture were used [1x buffer, 1.5 mM MgCl2, 0.25 of each dNTPs, 0.5 pmol/µL of each oligonucleotide (HDV-E and HDV-A) and 0.025 U/µL of Taq DNA polymerase (Invitrogen). The PCR conditions were: 94°C for 2 min, followed by 40 cycles of 30s at 94°C, 50s at 58°C and 45s at 72°C with a final extension of 5 min at 72°C. The PCR products were visualized in 1.5% agarose gel, amplified 403 bp and 374 bp respectively were obtained.^13,14^

### HDV sequencing

The amplified PCR products were purified with the QIAquick Gel Extraction Kit (QIAGEN, Hilden, Germany). The product was quantified using the nanodrop. Sequencing was performed with the oligonucleotides described above and the Big Dye Terminator v3.1 Cycle Sequencing kit (Applied Biosystems) on the ABI 3500 automated genetic analyzer (Applied Biosystem, Foster City, CA, USA).

### Phylogenetic Analysis

Phylogenetic analysis was performed with 111 reference sequences from the Genbank database (National Center for Biotechnology Information, Bethesda, MD, USA), corresponding to HDV genotypes 1 to 8. The 42 Peruvian sequences of the study were edited using the Chromas 2.6.5 program. To elaborate the consensus sequences, the Bioedit v7.2 program was used. The alignment of all the sequences was carried out with ClustalW v.1.6 of the Mega V7.0 program (https://www.megasoftware.net/), the cut was made with the Gblocks server software.

The Bayesian molecular analysis of the sequences was performed using the Markov Chain Monte Carlo (MCMC) of the Beast V2.5.2 program, with the GTR+G (General Time Reversible, gamma distributed) nucleotide substitution model obtained with the Jmodeltest v2.1.10. For the measurement of phylogenetic time, two clock models were used: relaxed uncorrelated lognormal molecular clock and relaxed uncorrelated Exponential molecular clock, with 30 million substitutions. The best molecular clock was chosen by Bayes Factor (BF) Comparison. Tree Annotator was used to obtain the tree with the maximum clade credibility (MCC) from the evaluation of each clade of the 30,000 trees excluding 10% of burn-in. The figtree v1.4.4 was used to visualize the phylogenetic tree. Beuti and Tracer v1.7 programs were also used.

### Evolutionary Analysis

For the evolutionary analysis, 47 reference sequences from GenBank corresponding to VHD-3 from South America from Brazil, Venezuela, Colombia, Bolivia and Peru, and the 42 sequences from this study were used. The Beast V2.5.2 software was used with the Bayesian Coalescing Exponential Poppulation model and the relaxed uncorrelated Exponential molecular clock, with 50 million substitutions and a burn-in of 10%. The nucleotide substitution model obtained in the Jmodeltest was HKY+G. The convergence of the chains was evaluated with the Tracer v1.7 software, the population dynamics over time was reconstructed with the Exponential Group Rate parametric model of the Coalecente Demographic Reconstruction, and with a high probabilistic density interval (HPD) at 95%.

### Statistic analysis

Demographic and virological information for each study sample was entered into an anonymous database in Excel 2013. Statistical analyzes were performed using STATA v.14.0 software (College Station, Texas) for Windows. The 95% CI was calculated for the study variables.

### Ethical considerations

The study protocol was approved by the ethics and research committee of the National Institute of Health (INS, Peru.

## RESULTS

Demographic and virological data of the 582 samples previously reported as positive for Anti-HBc by the ELISA method, 22.34% (130/582) corresponded to samples from residents of the province of Huanta, 13.40% (78/582) to the Kandozi ethnic group, 6.19% (N=36) to the Chapra ethnic group and 58.08% (338/582) to the Matsés ethnic group. Women represented 52.75% (N=307). The mean age was 38 years (0-86). Those younger than 5 years represented 0.74% (4/544) [95% CI, 0.20-1.87%] and those younger than 10 years represented 2.76% (15/544) [95% CI, 1.55-4.51%]. Of these samples, 17.35% (101/582) presented HBV-HDV co-infection, with 17.35% (95% CI, 14.36-20.68%) ELISA IgG-HDV positive and 9.11% (95% CI, 6.90-11.74 %) ELISA IgM-HDV positive 12.72% (74/582) were HBsAg positive. The viral load values for HBV were: undetectable in 01 sample, <2,000 IU/mL in 37 samples, and >2,000 IU/mL in 4 samples. Regarding origin, 4.95% (4/101) were from Huanta, 43.56% (44/101) from Kandozi, 31.68% (32/101) from Matsés and 19.80% (20/101) from Chapra.. Only 53.46% (54/101) were positive to nRT-PCR-HDV, the amplified fragment corresponding to nucleotides 857 to 1322, and they were used to determine the HDV genotype. Of the 50 amplified sequences, 42 sequences were considered for phylogenetic and evolutionary analysis. 8 sequences were not of good quality. (Table No. 1).

**Table 1.** Prevalence of HDV-IgG and HDV-IgM in the communities of the Amazon (Matsés, Kandozi and Chapra) and the inter-Andean valley of Huanta in reactive anti-HBc patients.

|  | N | (%) | HBsAG |  | ANTI-VHD (IgG/IgM) |  |
| --- | --- | --- | --- | --- | --- | --- |
|  |  |  | Positive |  | Positive |  |
|  |  |  | n (%) | IC 95% | n (%) | IC 95% |
| **All participants** | **582** | **100%** | **126 (21.65)** | **18.37-25.22** | **101 (17.35)** | **14.36-20.68** |
| **Gender** |  |  |  |  |  |  |
| Femenino | 307 | 52,75% | 56 (18.24) | 14.08-23.02 | 39 (12.70) | 9.19-16.95 |
| Masculino | 275 | 47,25% | 70 (25.45) | 20.41-31.03 | 62 (22.55) | 17.74-27.95 |
| **Age group (\*)** |  |  |  |  |  |  |
|  |  |  |  |  | (**) |  |
| 0 a 10 years | 15 | 2.76% | 0 (0,0) | 0,0-0,0 | 1 (6.67) | 0.17-31.95 |
| 11 a 18 | 27 | 4.96% | 9 (33.34) | 16.52-53.96 | 5 (18.52) | 6.30-38.08 |
| 19 a 29 | 143 | 26.3% | 55 (38.46) | 30.45-46.96 | 45 (31.47) | 23.97-39.76 |
| 30 a 59 | 297 | 54.60% | 53 (17.85) | 13.66-22.68 | 44 (14.81) | 10.98-19.37 |
| ≥ 60 | 62 | 11.40% | 5 (8.06) | 2.67-17.83 | 4 (6.45) | 1.79-15.70 |
| **Etnia** |  |  |  |  |  |  |
| Kandozi (ethnicity) | 78 | 13,40% | 42 (53.85) | 42.18-65.21 | 44 (56.41) | 44.70-67.61 |
| Chapra (ethnicity) | 36 | 6,19% | 22 (55.56) | 43.46-76.85 | 20 (55.56) | 38.10-72.06 |
| Matsés (Ethnicity) | 338 | 58,08% | 32 (9.46) | 6.57-13.10 | 32 (9.46) | 6.57-13.10 |
| Huanta (mestizo) | 130 | 22,34% | 30 (23,07) | 16.14-31.28 | 5 (3.87) | 1.26-8.75 |

\* The age group report includes 544 samples.
\*\* The age group report includes 99 samples (HBV-HDV co-infection)

### Nucleotide and amino acid analysis

The sequences obtained corresponded to the segment encoding aac 93 to 214 of the HDAg protein and to a non-translatable region at the C-terminal end. The percentage of Guanine and Cytosine bases was 62%. The percentage of similarity between the study sequences with the Peruvian sequence reported in 1993 was 94.0% to 99.71%.

Two of the RNA-binding domains (RDBs) were identified. The first domain is located between aac 96–106 (DQERRDHRRRK) and exhibits a change at aac R100Q. The second domain is between aac 135–145 (DDDERERRTAG) and shows changes in aac D135E, D137E and T143A in some of the analyzed sequences. Following the two RDB domains, there is a region rich in Proline and Glycine. The virus assembly signal (VAS) is located at the C-terminal end of HDAg-L, which corresponds to aac 196–214 (YGFTPPPPGYYWVPG-CTQQ), in some of the sequences analyzed a change was observed in the aac F198L, T199S and Y205H. The prenylation signal (Py), the composition of aac CTQQ and the poly(A) tail (952-TTTATT-957) did not present variation in any of the Peruvian sequences.

The target of RNA editing (nt1014) in most of the Peruvian sequences presents thymine (T), only 2 sequences from Chapra and 1 from Kandozi present 1014C. The RNA cleavage site for the antigenome is located at positions 904 and 905 (CG) and is constant in the Peruvian sequences obtained in the study and in the reference sequences used in the analysis.

### Phylogenetic and evolutionary analysis

The phylogenetic analysis of the 42 sequences obtained in this study and the 111 HDV sequences reported in GenBank showed that the sequences of the Matsés, Kandoshi, Shapra and Huanta communities are grouped in a monophyletic cluster with the sequences corresponding to genotype 3 that it is the most prevalent genotype in South America. (Figure No. 2)

**Figure 2.**
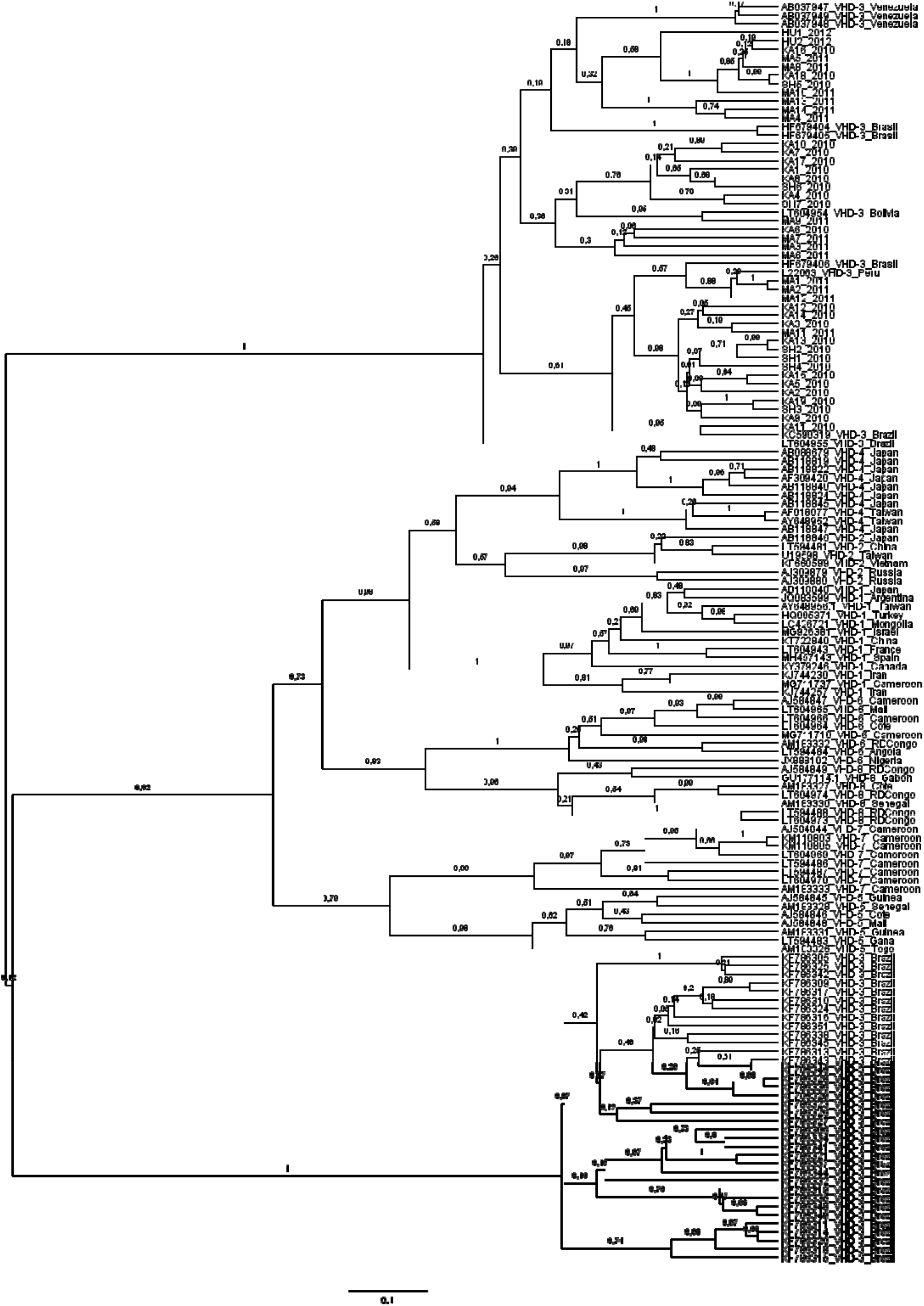
Unrooted HDV phylogenetic tree was estimated by Bayesian analysis of 153 hepatitis delta sequences. The 42 sequences obtained in this study are included. The genebank sequences are identified by their accession number and geographic origin.

The evolutionary analysis carried out using the Bayesian coalescent Exponential Population model for the 42 study sequences and the 47 HDV-3 sequences reported in GenBank allowed us to determine that the nucleotide substitution range of HDV-3 is 2047 10 -3 substitutions /site/year. The estimated value of the most recent common ancestor (tMRCA) for VHD-3 that has been circulating in South America is 1930 (HPD 95%). (Figure No. 3)

**Figure 3.**
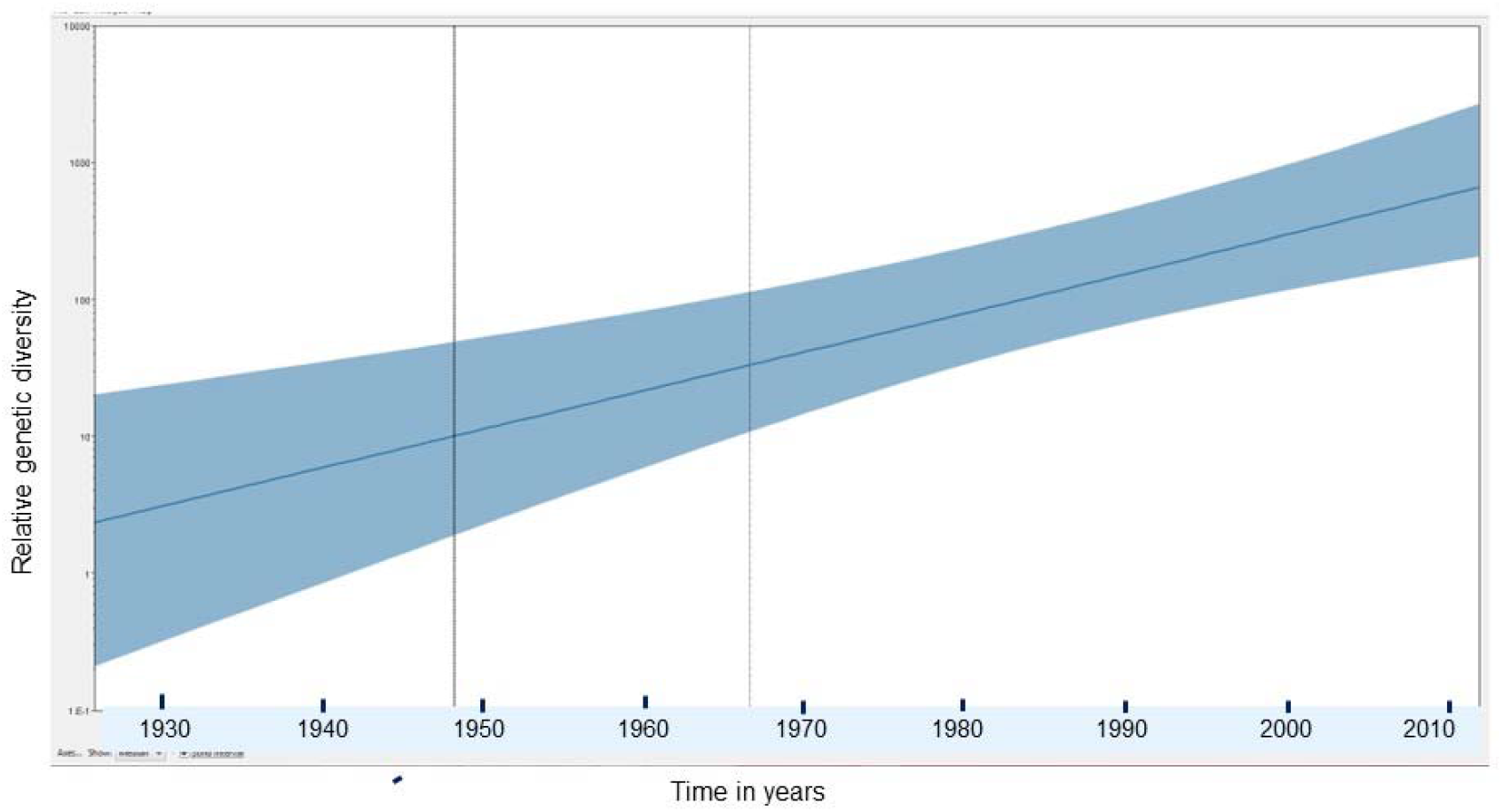
Population dynamics of the genetic diversity of HDV-3. The (y) axis represents the relative genetic diversity and the (x) axis the chronological time in years. The thick line in blue indicates the median and the thin lines the intervals of high probabilistic density (HPD) at 95% of the Ne.r. For the estimation, 89 HDV-3 sequences of 321 nt were used.

## DISCUSSION

In our study, we found the presence of HDV in La Huanta, an Andean region, and in the Amazonian communities of Matsés, Kandozi, and Chapra. Previous studies in Peru report the presence of HDV in some geographical areas such as Huanta, where a prevalence of 17.9% was found in HBV carriers^9^, while in Abancay the prevalence was 9% ^11^. In Andahuaylas and in the Amazon jungle, the prevalence of HDV in HBsAg carriers was 16.7% (8) and 39%, respectively^10^. Also in other Amazonian communities such as the Jíbaro, Pano and Arawak, a prevalence of 6.1% was found.

The high percentage of samples with HBV viral load values less than 2,000 IU/mL would be in agreement with studies that suggest that HDV suppresses HBV replication, producing low HBV viral load or below the confidence interval of the method^15,16^, although this requires a more precise evaluation of HBV variations.

The 42 sequences of the study have had a high percentage of similarity with the Peruvian sequence of 1993, which would indicate that these patients have probably been exposed to variants of HDV-3 that were closely related. Likewise, a high percentage of guanine and cytokine (62%) was observed, which agrees with the references that indicate that GC is more abundant than AT^17^ with percentages of 60% of GC^18^. The 19-amino acid sequence of HDAg-L is genotype-specific and varies between genotypes^19^. The ribozyme autocleavage site and RNA-binding domain are highly conserved. At the C-terminal end, the CTQQ motif has been found, which is the one conserved in the VHD-3 sequences, but it is very divergent for the aac T and Q in other genotypes^20^.

The presence of thymine (T) in nt 1014 is very frequent in genotype 3. The finding of 1014C in 3 Peruvian sequences is rare, there are only 2 similar reports in 1 sequence from Brazil and 1 from Bolivia (LT604954, LT604955). In other genotypes, the presence of 1014C is reported. This site is the position of heterogeneity that allows the formation of the 2 isoforms of HDAg by editing the RNA at the stop codon with the change of C for T to form tryptophan (aac195). The 1013-1015nt region corresponds to the end of the short-form Delta antigen protein (HDAg-S), which has a viral replication function. The 953-955nt region corresponds to the end of the long-form Delta antigen protein (HDAg-L) that promotes the HDV-RNA envelope for virion assembly ^21^.

The phylogenetic analysis allowed us to determine that the 42 sequences reported in this study correspond to HDV genotype 3, which is consistent with what was reported in 1993 in an outbreak of acute hepatitis in soldiers settled in four military posts in the Amazon jungle of Peru(12). This genotype has also been detected in Colombia^22^, Bolivia^23^, Brazil ^15,19^ and Venezuela^24,25^ and has been considered predominant in South America.

The range of VHD-3 nucleotide substitution obtained by Bayesian coalescent analysis was similar to those obtained by other authors who report ranges from 3 × 10 -2 to 3 × 10 -3 s/s/a (18), however It must be considered that these evolutionary ranges are not homogeneous throughout the genome, some consider 3.2 × 10 -3 s/s/a for the non-coding region and the coding region, 1.49 × 10 -3 s/s/a for non-coding. synonyms and 0.67 × 10 -3 s/s/a for synonyms^26^. The tMRCA has estimated that HDV-3 has been circulating in South America since 1930. There are reports of the presence of HDV infection since 1934 in the Amazon region ^27^.

## Data Availability

All data produced in the present study are available upon reasonable request to the authors

## Author Contribution

CACS, JNBT, LSS, and RDNO contributed to study design, data collection, data analysis, and approval of the final version of the article. CACS and JNBT contributed to the writing of the article. LSS contributed to the processing of the samples. JNBT contributed to the bioinformatic analysis. All authors agree with the final version of the article.

## Acknowledgment

The authors thank the staff of the National Institute of Health - National Hepatitis Reference Laboratory for providing the facilities for the execution of this project.

